# An explainable, bias-audited machine learning model to detect hidden hepatitis C from region-wide real-world health records

**DOI:** 10.64898/2026.11.05.26367298

**Authors:** Alberto Esteban-Medina, Carlos Loucera, Joaquín Dopazo, Manuel Romero Gómez

## Abstract

**Background:** Although direct-acting antivirals have transformed hepatitis C virus (HCV) treatment, 25-30% of active infections in Spain remain undiagnosed. We aimed to develop and validate an interpretable machine-learning model to identify undiagnosed HCV infection using only data already recorded within the health system.

**Materials and Methods:** We conducted a retrospective, population-based cohort study using the Andalusian Population Health Database (BPS). Individuals diagnosed with HCV between 2017 and 2024 (n=25,028) were matched, up to 10:1, to controls by birth year, sex, and health district. An Explainable Boosting Machine was trained on 2017-2022 data, with a threshold set in a 2023 validation set to guarantee ≥60% sensitivity, then evaluated unmodified in an independent, non-matched, population-representative 2024 sample approximating real-world screening. Predictors comprised chronic-disease diagnoses and laboratory summary statistics recorded ≥30 days before the index date; age and sex were excluded as matching variables. Predictors reflecting socioeconomic proxies or matching artefacts (pharmaceutical co-payment category, a zone-level deprivation index) were removed after explainability review.

**Results:** In the independent test set, the model achieved an AUC of 0.837 (0.826 in validation), sensitivity 63.5%, specificity 91.8%, positive predictive value 7.0%, and negative predictive value 99.6%. Leading predictors were minimum alanine aminotransferase, HIV infection, dependence on non-tobacco substances, and platelet-count statistics, forming a clinically coherent, laboratory- and comorbidity-driven profile.

**Conclusion:** An explainable model built exclusively from routine electronic health record data can identify undiagnosed HCV infection with discrimination comparable to previously published tools, evaluated under conditions approximating true population prevalence. Bias-auditing was feasible without sacrificing interpretability, supporting a scalable, equity-conscious instrument for the final phase of HCV elimination.

## Introduction

Hepatitis C virus (HCV) infection remains one of the principal preventable causes of chronic liver disease, cirrhosis, and hepatocellular carcinoma worldwide (Gamkrelidze et al., 2021). The introduction of direct-acting antivirals (DAA) after 2015 transformed the therapeutic landscape, achieving cure rates above 95% (Zhang, 2016) and prompting the World Health Organization (WHO) to establish the goal of eliminating viral hepatitis as a public health threat by 2030, including diagnosis of 90% of infected individuals and a 90% reduction in incidence (WHO, 2022). As therapeutic effectiveness is no longer the principal limiting factor, case finding has become one of the central bottlenecks in the final phase of HCV elimination.

Spain is internationally recognized as one of the countries closest to these targets with the introduction of universal access to direct-acting antivirals and the implementation of the Strategic Plan for Tackling Hepatitis C, in 2015. Since then, more than 172,000 patients have been treated with DAA, reducing the estimated national prevalence of active infection to approximately 0.14% (del Amo Valero et al., 2025; Romero Otero et al., 2026). Despite this progress, an estimated 25-30% of active infections remain undiagnosed, a proportion that has changed little in absolute terms since the baseline national seroprevalence survey of 2018, which already reported an undiagnosed fraction of 29.4% (Estirado Gómez et al., 2021; Thomadakis et al., 2026). These remaining infections are increasingly concentrated among people who are not reached efficiently by conventional pathways targeting individuals under classical risk factors, including those with remote exposures, people who inject drugs, migrants from higher-prevalence regions, people with a history of incarceration, patients with HIV or other blood-borne infections, and socially vulnerable populations. Some of these individuals nevertheless interact repeatedly with primary care, emergency departments, hospitals, laboratories or pharmacy services, leaving potentially informative clinical traces before HCV is suspected. This hidden reservoir is now regarded as the principal obstacle to elimination, since residual transmission and disease burden concentrate among individuals outside the conventional healthcare circuit or who have never been tested (Romero Otero et al., 2026).

Beyond conventional screening, or opportunistic screening in selected healthcare settings, simple clinical risk scores can improve targeting. For example, Structured questionnaires and simple risk scores developed for primary care (Martínez-Sanz et al., 2022; McGinn et al., 2008; Nguyen et al., 2005) achieve reasonable discrimination (areas under the curve typically between 0.77 and 0.90), but depend on the availability, completeness, and truthfulness of self-reported risk factors, which are inconsistently captured in daily practice. The increasing availability of longitudinal electronic health records (EHRs), administrative claims, laboratory data, prescriptions and healthcare-utilisation histories provides an alternative strategy: identifying high-risk individuals using only information already generated during routine care. Early large-scale studies using US insurance claims showed that ensemble models could achieve substantially higher precision than universal or risk-based screening at equivalent recall (Doyle et al., 2020), a finding later replicated with gradient-boosting models trained on ambulatory electronic medical records (Rigg et al., 2023). Further studies extended this approach by complementing structured EHR variables with risk factors extracted from clinical narratives through natural language processing (Fong et al., 2023; Sánchez-Antolín et al., 2026). Large databases produced models with excellent discrimination for high-risk HCV infection among tested individuals (Jang et al., 2025), and some RWD-based models were prospectively deployed in the health system (Dagan et al., 2024). Primary-care data from almost two million patients in North-East London also showed that HCV was the most accurately predicted of three blood-borne viruses using routinely recorded demographic, clinical and behavioural indicators (Manley et al., 2026).

Despite these advances, a recent systematic review identified only 13 HCV prediction models and found substantial heterogeneity, high risk of bias in most studies, scarce primary-care validation and inconsistent reporting (Leber et al., 2025). Many models are trained only among tested patients or use a future diagnosis as a proxy for true infection, creating verification and label bias. Other concerns include temporal leakage, class imbalance, low positive predictive value at population prevalence, limited calibration, and uncertain transportability across health systems and epidemiological contexts.

The present study aimed to develop and validate an interpretable predictor of previously undiagnosed or hidden HCV infection using exclusively routinely collected longitudinal data available within the health system. By identifying individuals with a high probability of infection without requiring questionnaires, new examinations or additional data collection, this approach seeks to complement existing screening strategies and provide a scalable instrument for the final phase of HCV elimination.

## Material and methods

### Study design, setting and data source

We conducted a retrospective, population-based cohort study to develop and temporally validate a machine-learning model for the identification of undiagnosed (hidden) HCV infection using data routinely collected within the public health system of Andalusia (Southern Spain). Clinical information was requested to the “Base Poblacional de Salud de Andalucía” (BPS) (Muñoyerro-Muñiz et al., 2020), a regional repository that integrates electronic health records, laboratory results, and diagnostic coding for more than 16 million users of the Andalusian Health system, under the existing regional framework for secure secondary use of health data (Muñoyerro-Muñiz et al., 2025).

### Study population

All individuals diagnosed with HCV infection in the BPS between 2017 and 2024 were identified as cases. A control cohort up to 10 times larger than the case cohort was selected from the same source population, matched by sex, age, and health district. Matched controls were assigned the same index date as their corresponding case (the date of HCV diagnosis).

In addition, an independent sample of randomly selected controls was drawn from the BPS, selected independently of the matched case-control dataset and not matched to any individual case. Eligibility for this independent control group required that individuals be alive throughout 2017–2023. Each was assigned a random index date within 2024.

### Clinical Data

Clinical data corresponding to the patients meeting the inclusion criteria was transferred from BPS to the Platform of Generation of Medical Evidence (PAGEM), a Trusted Research Environment for secondary use of clinical data (Muñoyerro-Muñiz et al., 2025).

All clinical data available in the electronic health record from 30 days prior to the index date to the end of the patient’s record, without any additional questionnaire or dedicated data collection, will be used to build the predictor model. Two domains of variables were considered: (i) chronic diseases and diagnostic history, including hypertension, diabetes, obesity, tobacco dependence, dependence on other substances, chronic liver disease other than cirrhosis, and HIV infection; and (ii) clinical laboratory analytes recorded at any point before the index date (including haemoglobin, alanine aminotransferase, aspartate aminotransferase, gamma-glutamyl transferase, platelet count, and total protein, among others), summarised for each patient using the mean, minimum, maximum, and standard deviation of all available determinations.

### Data partitioning and threshold selection

The case-control sample was partitioned into training, validation, and test sets using a temporal, non-overlapping criterion based on the year of HCV diagnosis, so that the model was trained exclusively on historical cases and evaluated on progressively more recent, unseen periods. The training set comprised diagnoses from 2017–2022, the validation set comprised diagnoses from 2023, and the test set comprised diagnoses from 2024.

Two complementary modelling strategies were evaluated. The first included all available features, whereas the second excluded the income-based pharmaceutical copayment bracket and the area-level socioeconomic index. The income-based pharmaceutical copayment bracket and the area-level index were removed to reduce the risk of the model primarily capturing socioeconomic disadvantage rather than clinically relevant risk.

For each individual, a single prediction was generated 30 days before the corresponding index date, using only the clinical information available up to that time point. The index date was defined as the diagnosis date for cases, the matched date for matched controls, and a randomly assigned date for controls in the 2024 test set. This approach ensured that predictions reflected only information available before the relevant reference date and prevented any post-diagnosis information from contributing to the risk score.

The classification threshold was selected exclusively in the validation set, by requiring a minimum sensitivity (true positive rate) of 60%, prioritising case detection as appropriate for a screening application. This threshold, together with the aggregation function from which it was derived, was then applied without modification to the independent test set to obtain an unbiased estimate of performance.

### Prediction horizon and observation window

To reduce the risk of information leakage and minimise bias related to diagnostic work-up immediately preceding hepatitis C diagnosis, a prediction horizon was established 30 days before the index date. For each individual, only clinical data recorded before this time point was used to generate predictor variables.

The observation window therefore included all available clinical information prior to 30 days before the index date. This approach was intended to ensure that the model relied on pre-existing clinical information rather than on tests, diagnoses, or healthcare encounters directly associated with the eventual diagnosis of hepatitis C. Alternative prediction horizons were explored in sensitivity analyses to evaluate their impact on model performance.

### Feature selection and feature engineering

Candidate predictors were derived from two main sources of clinical information: chronic disease diagnoses and laboratory test results. Chronic conditions were selected from the information available in the BPS, while laboratory variables were chosen based on their clinical relevance after review by expert hepatologists.

Chronic diseases were encoded as binary variables, indicating whether each condition was present or absent before the prediction horizon. Laboratory results were processed to capture both the central tendency and longitudinal variability of each biomarker over time. For each selected laboratory parameter, the following summary features were generated using all available values recorded before the prediction horizon: mean, standard deviation, minimum value, and maximum value.

Sex and age were not included as predictors in the model because they had already been used as matching variables in the matched case-control design. This decision was made to reduce the possibility that the model would learn patterns driven by the matching procedure rather than clinically meaningful predictors of hepatitis C infection.

### Model development

Predictive models were developed using an Explainable Boosting Machine (EBM), a generalized-additive-model-based gradient boosting algorithm that combines predictive performance comparable to standard black-box ensembles with feature-level interpretability, allowing the relative contribution of each predictor to the estimated risk to be quantified (Caruana et al., 2015; Nori et al., 2019).

For patients diagnosed with HCV, only information recorded before their individual diagnosis date was used for feature construction. For controls, all information available up to their matching date was used.

### Threshold selection

The classification threshold was selected in the validation set by requiring a minimum sensitivity (true positive rate) of 60%, prioritising case detection as appropriate for a screening application, and the same threshold was subsequently applied, without modification, to the independent test set.

### Outcome metrics

Model performance was assessed using sensitivity (true positive rate), specificity (true negative rate), false positive rate, positive predictive value, negative predictive value, F1-score, Matthews correlation coefficient, and the false discovery and false negative rates, together with receiver operating characteristic (ROC) curves for the training, validation, and test sets.

## Results

### Study population

A total of 25,028 individuals diagnosed with hepatitis C virus (HCV) infection between 2017 and 2024 were identified, together with 213,404 matched controls used for training and validation (a realized ratio of approximately 9.2 controls per case, slightly below the targeted 10:1 ratio due to pairs excluded for incomplete matching data), and 193,054 independent, non-matched controls used for the final test evaluation (Table 1). The training set comprised 21,008 cases and 194,112 matched controls with an index date between 2017 and 2022 and the validation set comprised 2,139 cases and 19,292 matched controls with an index date in 2023. The test set comprised 1,881 cases diagnosed in 2024, evaluated against the independent random control sample of 193,054 individuals rather than their matched counterparts, to approximate population-level screening conditions.

**Table 1.**
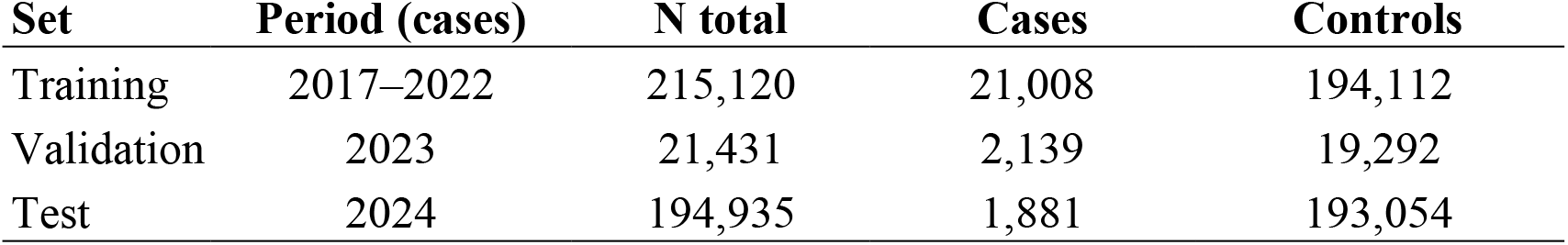
Distribution of cases and controls across the training, validation, and test sets. Test-set cases were evaluated against an independent, non-matched sample

Baseline characteristics of the three groups are summarised in Supplementary Table 1. Matching performed as intended: cases and matched controls were nearly identical in sex (male, 67.9% vs. 68.4%) and age (mean 59.5 [SD 13.3] vs. 59.7 [13.0] years), whereas the independent random control sample, drawn without matching, was younger and more balanced by sex (49.9 [17.9] years, 48.6% male), as expected for a population-representative comparator. Relative to matched controls, the conditions most strongly over-represented among cases were those that characterise the known epidemiology of HCV: HIV infection (14.0% vs. 0.4%), dependence on substances other than tobacco (15.7% vs. 0.9%), alcohol dependence (16.7% vs. 4.2%), tobacco dependence (23.0% vs. 11.5%), chronic hepatitis B without delta-agent (1.4% vs. 0.2%), and the full spectrum of chronic liver disease, including unspecified cirrhosis of liver (8.3% vs. 0.2%), liver cirrhosis (5.2% vs. 0.2%), chronic liver disease except cirrhosis (5.6% vs. 0.5%), portal hypertension (1.6% vs. 0.1%), and liver and biliary tract cancer (3.0% vs. 0.1%). Mental-health and neuropsychiatric diagnoses were also more frequent among cases (schizophrenic disorder 5.8% vs. 1.4%; adult personality and behaviour disorder 5.4% vs. 1.2%; other organic mental disorder 7.1% vs. 2.8%; mood disorder 9.9% vs. 5.9%; anxiety disorder 30.4% vs. 21.2%), as were chronic obstructive pulmonary disease (15.8% vs. 7.9%), chronic kidney failure (5.5% vs. 2.6%), and heart failure (8.8% vs. 5.1%). By contrast, several highly prevalent cardiometabolic and musculoskeletal conditions were comparable between the two groups or less frequent among cases (hypertension 39.2% vs. 40.7%; osteoarthritis and spondylosis 27.1% vs. 28.3%; obesity 6.8% vs. 9.2%; gout and other crystal arthropathies 3.1% vs. 4.7%), indicating that the comorbidity profile separating cases from controls is concentrated in blood-borne co-infection, substance use, and liver disease rather than in general chronic-disease burden. Differences with respect to the random control sample point in the same direction but are additionally influenced by the age and sex imbalance described above, since this group was not matched to cases.

### Model discrimination

Using the 30-day prediction horizon, the model achieved an AUC of 0.841 in the training set, 0.826 in the validation set, and 0.837 in the independent test set (Figure 1A), indicating that discrimination was preserved when the model was evaluated against a population-representative, non-matched control sample rather than matched controls. The corresponding area under the precision-recall curve (AUPRC) was 0.591 (training), 0.542 (validation), and 0.240 (test) (Figure 1B). The marked reduction in test-set AUPRC reflects the much lower case prevalence in this dataset: the dashed reference line in Figure 1B (0.058) corresponds to the pooled case prevalence across the combined training, validation, and test datasets, whereas the true prevalence within the test set alone was considerably lower, at approximately 1.0% (1,881 cases among 194,935 individuals). Relative to this true test-set prevalence, the observed AUPRC of 0.240 represents a substantially larger enrichment over chance than a direct comparison with the plotted baseline would suggest.

**Figure 1.**
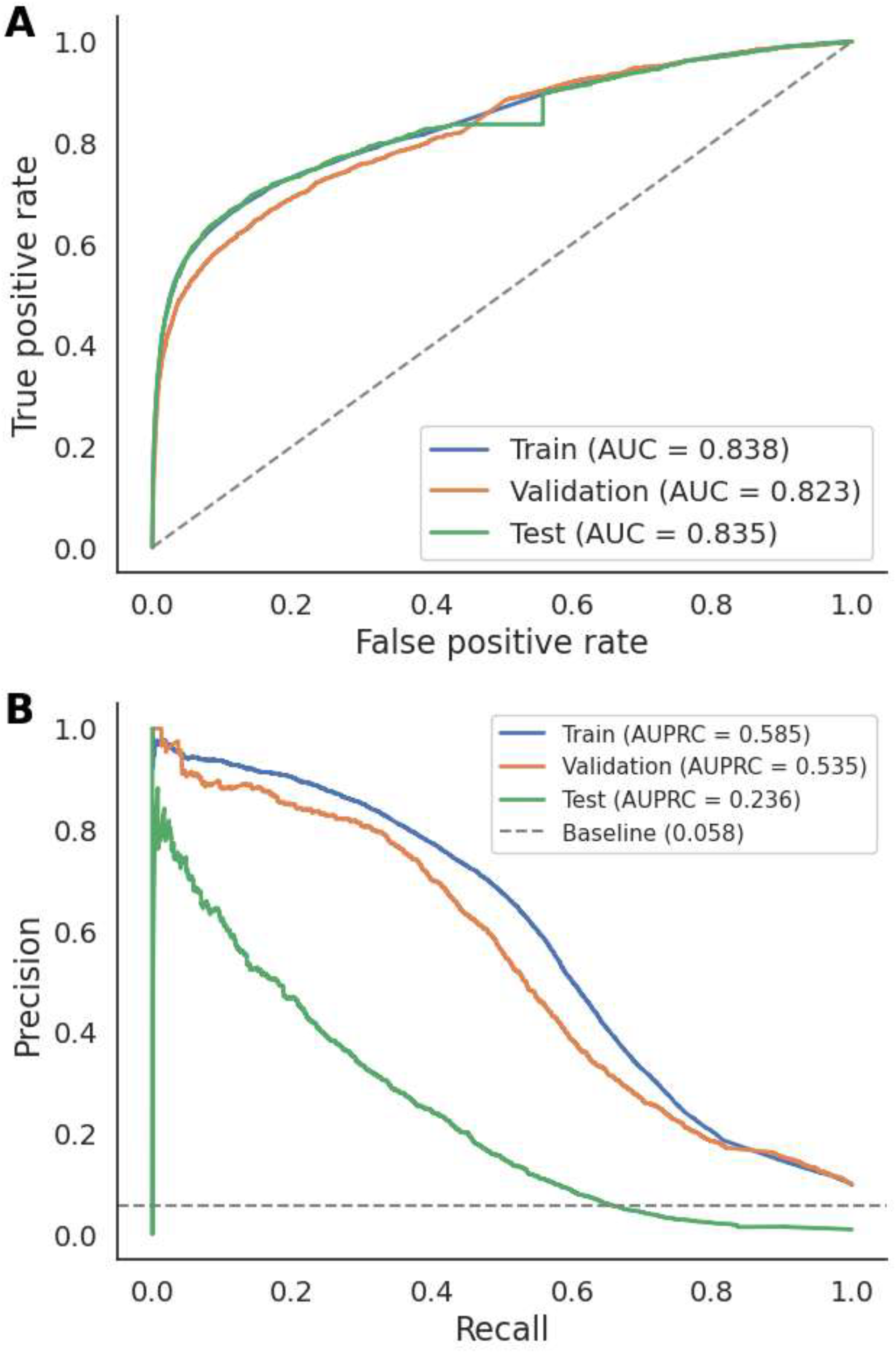
Model discrimination in the training, validation, and test sets, using the 30-day prediction horizon. (A) Receiver operating characteristic (ROC) curves. AUC, area under the ROC curve. (B) Precision-recall curves. AUPRC, area under the precision-recall curve. The dashed line in panel B indicates the pooled case prevalence across the combined training, validation, and test datasets (5.8%), provided as a general reference rather than a dataset-specific baseline.

### Classification performance in the test set

A classification threshold of 0.1793 was selected in the validation set to guarantee a minimum sensitivity of 60% and applied without modification to the test set. At this threshold, the model correctly classified 1,195 of 1,881 true cases (sensitivity 63.5%) and 177,167 of 193,054 true controls (specificity 91.8%) (Table 2). Table 3 summarises the corresponding classification metrics. Consistent with the low prevalence of active HCV infection in this population-representative test set, precision (positive predictive value) was low (7.0%), meaning that most individuals flagged by the model above threshold would not, in fact, have undiagnosed HCV infection; the negative predictive value, by contrast, was high (99.6%), indicating that a prediction below threshold reliably identified individuals unlikely to be infected.

**Table 2.**
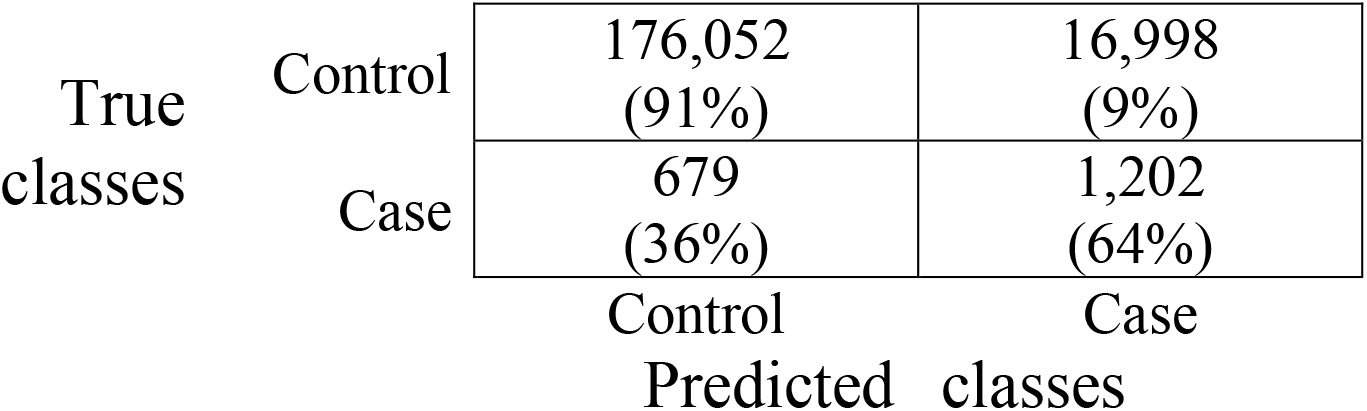
Confusion matrix for the test set, at the classification threshold selected in validation to guarantee a minimum sensitivity of 60%. Cell values show the number of individuals and, in parentheses, the row-wise percentage (i.e., relative to the true class).

|  |  | Predicted classes |  |
|---|---|---|---|
|  |  | Control | Case |
| True classes | Control | 176,052<br>(91%) | 16,998<br>(9%) |
|  | Case | 679<br>(36%) | 1,202<br>(64%) |

**Table 3.**
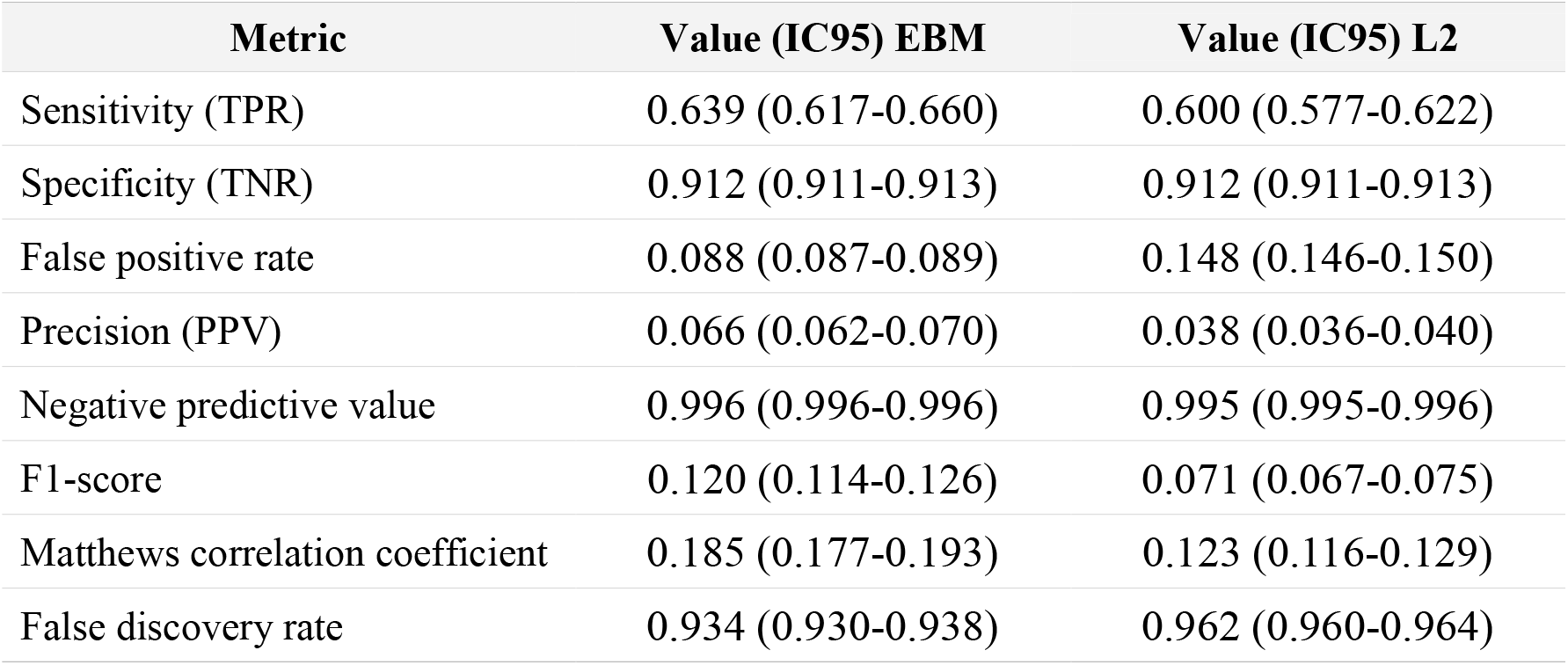
Classification performance in the test set at the validation-derived threshold EBM (Explainable Boosting Machine) vs L2 (Logistic Regression).

| **Metric** | **Value (IC95) EBM** | **Value (IC95) L2** |
|---|---|---|
| Sensitivity (TPR) | 0.639 (0.617-0.660) | 0.600 (0.577-0.622) |
| Specificity (TNR) | 0.912 (0.911-0.913) | 0.912 (0.911-0.913) |
| False positive rate | 0.088 (0.087-0.089) | 0.148 (0.146-0.150) |
| Precision (PPV) | 0.066 (0.062-0.070) | 0.038 (0.036-0.040) |
| Negative predictive value | 0.996 (0.996-0.996) | 0.995 (0.995-0.996) |
| F1-score | 0.120 (0.114-0.126) | 0.071 (0.067-0.075) |
| Matthews correlation coefficient | 0.185 (0.177-0.193) | 0.123 (0.116-0.129) |
| False discovery rate | 0.934 (0.930-0.938) | 0.962 (0.960-0.964) |

Based on these results, the maximum-probability aggregation function was retained for both models, as it achieved an equivalent or lower false positive rate than the minimum function at the same sensitivity target. The full-population model with maximum aggregation was selected as the primary model, and its corresponding threshold (0.1437) was fixed for the blind evaluation in the test set. The remaining three model/function combinations were retained for comparison.

Figure 2 shows the 30 most important predictors of the reduced model (i.e., excluding pharmaceutical co-payment category and the zone-level deprivation index). The minimum recorded alanine aminotransferase (ALT) value was the single strongest predictor, followed by HIV infection, dependence on substances other than tobacco, and summary statistics of platelet count. Beyond these leading variables, the ranking included obesity, aspartate aminotransferase (AST), chronic obstructive pulmonary disease, anxiety disorder, hypertension, unspecified cirrhosis and other unspecified liver disease, haemoglobin, prothrombin time, gout and other crystal arthropathies, tobacco dependence, C-reactive protein, alkaline phosphatase, total protein, and gamma-glutamyl transferase (GGT), a predictor set dominated by liver-related and haematological laboratory markers together with a small number of comorbidities and substance-use diagnoses, and notably free of the age-, sex-, and matching-related variables excluded by design. The corresponding shape functions, which describe how the contribution to the model score varies across the range of each predictor, are shown for the 15 most important variables in Supplementary Figure 3 and for those ranked 16th to 30th in Supplementary Figure 4.

**Figure 2.**
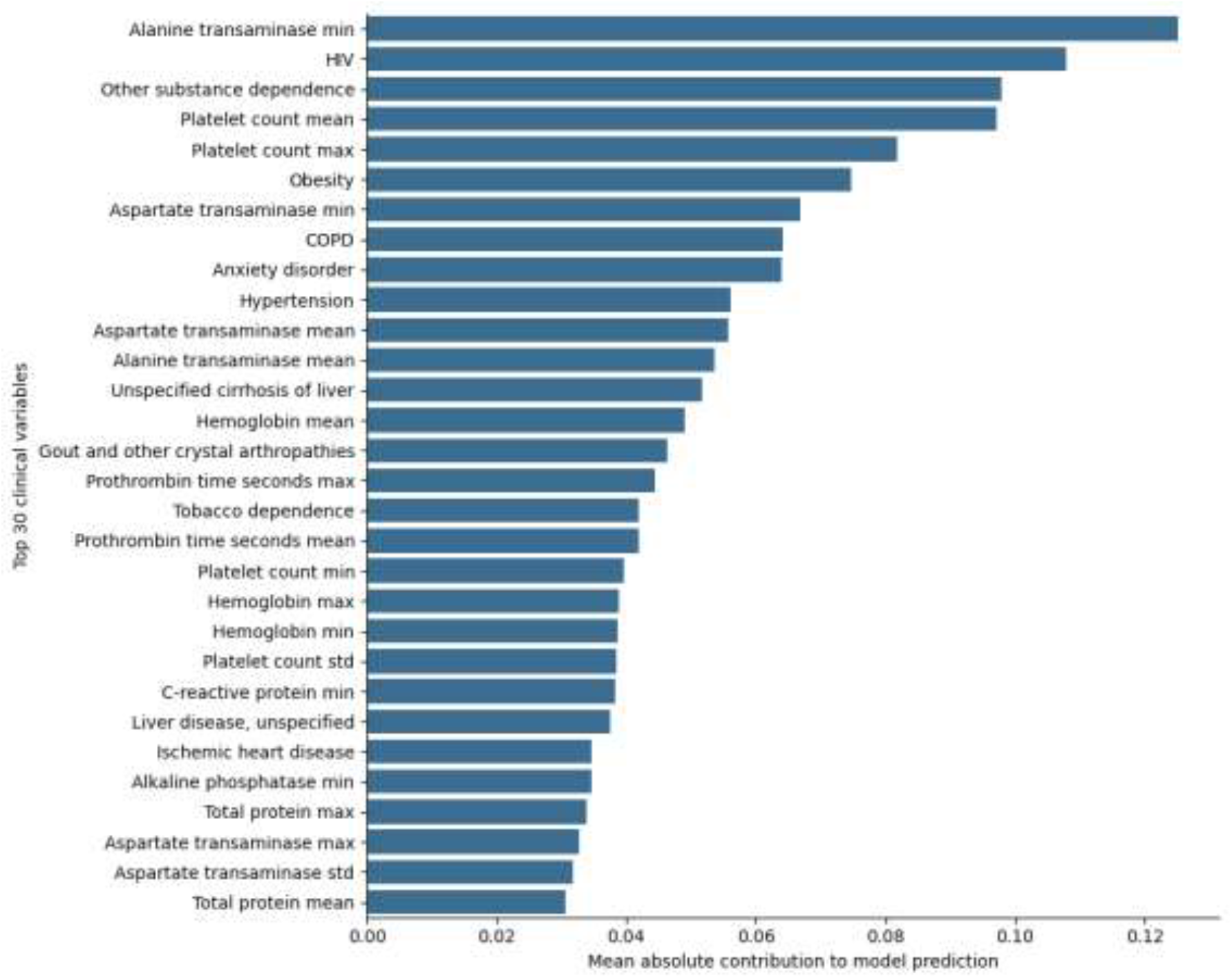
Predictor importance for the reduced model (excluding pharmaceutical co-payment category, zone-level deprivation index, and dyslipidaemia). Bars represent the mean absolute contribution of each variable to the model’s predicted risk score, ranked in descending order for the 30 highest-ranked predictors.

**Figure 3.**
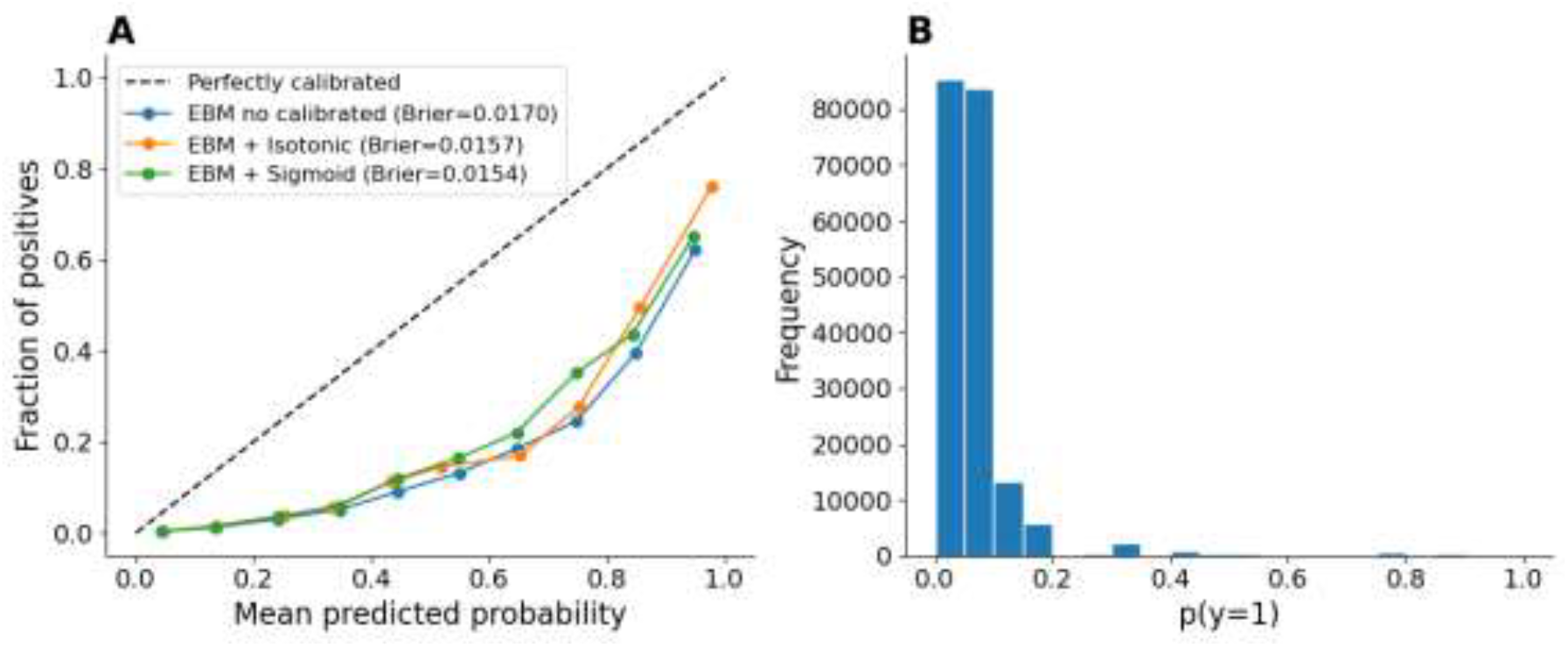
Model calibration in the test set. (A) Reliability diagram comparing the uncalibrated Explainable Boosting Machine (EBM) with isotonic- and sigmoid-calibrated versions. Brier scores are shown in the legend. (B) Distribution of predicted probabilities of hepatitis C infection across the test population.

### Variable relevance for the prediction

Figure 2 shows the 30 most important predictors of the reduced model (i.e., excluding pharmaceutical co-payment category, the zone-level deprivation index, and dyslipidaemia). The minimum recorded alanine aminotransferase (ALT) value was the single strongest predictor, followed by HIV infection, dependence on substances other than tobacco, and summary statistics of platelet count. Beyond these leading variables, the ranking included obesity, aspartate aminotransferase (AST), chronic obstructive pulmonary disease, anxiety disorder, hypertension, unspecified cirrhosis and other unspecified liver disease, haemoglobin, prothrombin time, gout and other crystal arthropathies, tobacco dependence, C-reactive protein, alkaline phosphatase, total protein, and gamma-glutamyl transferase (GGT) — a predictor set dominated by liver-related and haematological laboratory markers together with a small number of comorbidities and substance-use diagnoses, and notably free of the age-, sex-, and matching-related variables excluded by design.

### Sensitivity analyses

Two pre-specified sensitivity analyses were conducted. First, the prediction horizon was varied across nine values (30, 60, 90, 120, 160, 180, 270, 360, and 720 days before the index date). Model performance declined progressively as the horizon was extended, supporting the choice of the 30-day horizon for the primary analysis (classification metrics at the validation-derived threshold for every horizon are shown in Supplementary Figure 1, and the corresponding AUROC and AUPRC values in Supplementary Figure 2). Second, a full-variable model retaining the pharmaceutical co-payment category and the zone-level deprivation index was trained and evaluated in parallel with the reduced model; the corresponding results are reported in Supplementary Table 2, since the reduced model was adopted as the primary analysis.

### Calibration

Because the classification threshold approach required a single decision criterion for the confusion matrix in Table 2, formal probability calibration (isotonic regression and Platt/sigmoid scaling, both fitted on the validation set) was explored as an alternative but not adopted for the primary analysis. Calibration modestly improved the Brier score in the test set (uncalibrated EBM, 0.0169; isotonic calibration, 0.0156; sigmoid calibration, 0.0154), but reliability diagrams showed that all three approaches underestimated the true fraction of positives at higher predicted probabilities, without a clear advantage for either calibration method (Figure 3A). The distribution of predicted probabilities was strongly right skewed, with most individuals assigned a low predicted risk, consistent with the low prevalence of active HCV infection in the test population (Figure 3B).

## Discussion

In this population-based study of the entire Andalusian health system, a machine-learning model built exclusively from structured electronic health record data (including chronic disease diagnoses and longitudinal laboratory summary statistics, without any self-reported risk factor) identified 63.5% of patients who would go on to receive an HCV diagnosis in 2024, at a specificity of 91.8%, with an AUC of 0.837 in this fully independent, population-representative test set (compared with 0.826 in validation and 0.841 in training), indicating that discrimination was well preserved despite the marked change in case-mix between the matched development cohorts and the unmatched test population. Precision at this operating point was low (7.0%), a direct and expected consequence of evaluating the model against a control population with a true HCV prevalence of approximately 1%, rather than the artificially enriched prevalence of a matched case-control sample. The negative predictive value, conversely, was high (99.6%). Predictor importance was dominated by longitudinal liver-related and haematological laboratory markers (alanine aminotransferase, aspartate aminotransferase, platelet count) together with HIV infection, substance dependence, obesity, and a small number of chronic comorbidities, a clinically coherent set of variables, once age, sex, and two variables identified as artefacts of the matched design (two socioeconomic proxies) were removed.

These results presented here extend a rapidly growing body of work applying machine learning to routinely collected health-system data for HCV case-finding. The most directly comparable efforts used US insurance claims (Doyle et al., 2020) and ambulatory electronic medical records (Rigg et al., 2023), respectively, and reported their models’ advantage as a fold-improvement in precision over risk-based or universal screening rather than as a conventional AUC, limiting direct numerical comparison. Studies reporting a standard discrimination statistic offer a closer benchmark. The use of structured EHR data in a US academic health system achieved a C-statistic of 0.777 (Fong et al., 2023), while a value of 0.916 was reported using a larger all-payer claims database (Jang et al., 2025). In a prospective, population-wide deployment within an integrated Israeli health system, an AUROC of approximately 0.95 was reported together with a number needed to screen (NNS) of 10, versus 1,029 for conventional risk-based screening (a roughly 100-fold efficiency gain) (Dagan et al., 2024). Recently, a review of published HIV, hepatitis B, and HCV prediction models, reported a pooled AUC of 0.80 across 13 comparable HCV prediction models (Leber et al., 2025).

Our results sit at or above this pooled estimate but were obtained under a design explicitly built to avoid several of the specific weaknesses (matched or convenience test sets, absence of calibration assessment, unclear temporal separation between development and evaluation data) observed in most of the published predictors, as highlighted in the review (Leber et al., 2025).

Within Spain, the most direct comparator remains the LiverTAI model (Sánchez-Antolín et al., 2026), which combined electronic health records with natural language processing across six hospitals to detect unconfirmed HCV infection, reporting an AUC of 0.794 on independent geographic validation. Our model achieved higher discrimination (AUC 0.837 vs. 0.794) than LiverTAI, using structured data alone, without a natural-language-processing component, although the two studies target related but distinct outcomes (undiagnosed active infection versus unconfirmed serology) and are not strictly interchangeable. Our findings also complement two other Spanish initiatives built on automated, laboratory-triggered case detection rather than statistical risk modelling, Intelligen-C (Castro Urda et al., 2025) and CriVHado C (Griñó et al., 2025), illustrating two complementary approaches (rule-based alerting versus continuous risk scoring) to the same underlying public-health problem of residual undiagnosed infection, estimated at 25-30% of active cases nationally despite treatment coverage exceeding 80% of diagnosed patients (Romero Otero et al., 2026), against the WHO’s 2030 elimination targets (Gamkrelidze et al., 2021).

Compared with earlier, questionnaire-based risk-assessment tools like self-administered instrument (Nguyen et al., 2005), primary-care screening tool (AUC 0.77) (McGinn et al., 2008), and the five-item score (C-statistic 0.896) (Martínez-Sanz et al., 2022) our model’s discrimination is comparable to or below the latter. The distinction, however, is one of implementation rather than accuracy alone: those instruments require prospectively administered questionnaires capturing self-reported risk behaviour at the point of care, whereas our model operates entirely on data already present in the health record, requiring no patient contact, no clinician time, and no additional data-collection infrastructure, and can in principle be applied retrospectively across an entire historical population rather than only to patients presenting for a specific encounter.

Regarding the findings that our explainable approach has, the prominence of transaminases, platelet count, and other liver-related and haematological markers as relevant variables for the classification is consistent with the biochemical abnormalities long used as opportunistic screening triggers in Spain and elsewhere (Romero Otero et al., 2026), while the appearance of HIV infection and dependence on substances other than tobacco among the leading predictors mirrors the well-documented concentration of incident and recently acquired HCV infection among people living with HIV who report chemsex-associated substance use, particularly among men who have sex with men (Martín-Carbonero et al., 2024). The presence of obesity, hypertension, and markers of hepatic and haematological dysfunction (unspecified cirrhosis, prothrombin time, other unspecified liver disease) further aligns with a population accumulating undiagnosed liver injury before eventual diagnosis, of the kind identified through opportunistic testing in Spanish emergency departments (Camelo-Castillo et al., 2024). The final predictor set, obtained after removing pharmaceutical co-payment category, and a zone-level deprivation index, is free of variables that had shown implausible or bias-prone associations with the outcome, and is, correspondingly, more directly interpretable in clinical and biological terms. This pattern of laboratory- and comorbidity-driven prediction, rather than one dominated by demographic or socioeconomic proxies, is also broadly consistent with the determinants identified for HIV, HCV, and HBV risk in a comparable primary-care machine-learning model (Manley et al., 2026).

This study presents several characteristics that, taken together, distinguish it from previously published HCV case-finding models. First, as before, the analysis draws on a genuinely population-scale, multi-year cohort (more than 430,000 individuals across an entire regional health system followed from 2017 to 2024) evaluated under a strict, non-overlapping temporal split. Second, and most distinctively, the final test evaluation was deliberately conducted against an independent, non-matched, population-representative control sample rather than the matched controls used for model development. This is a design choice that directly addresses the verification bias and limited real-world validity (Leber et al., 2025) a drawback of most published models in this field, and which, to our knowledge, few comparable HCV prediction studies have explicitly implemented. Third, the exclusion of pharmaceutical co-payment category, and a zone-level deprivation index was not an incidental modelling choice but the outcome of a deliberate bias-auditing step, in which model explainability was used prospectively to detect and remove predictors likely to encode socioeconomic proxies or artefacts of the matched design rather than genuine clinical signal. This is an equity-oriented methodological safeguard that is largely absent from the prior literature reviewed here. Fourth, the systematic evaluation of nine prediction horizons spanning 30 to 720 days before diagnosis provides, to our knowledge, the most granular published assessment of how model performance decays with increasing lead time, information directly relevant to designing a realistic screening workflow rather than a single, retrospectively optimal cut-point. Fifth, formal calibration was assessed and reported transparently, including its limitations, rather than assumed from discrimination alone, a step recommended but rarely implemented in this literature. Finally, retaining the EBM architecture (Caruana et al., 2015; Nori et al., 2019) preserved full interpretability of the final predictor set throughout this bias-auditing process, which would not have been straightforward with a conventional black-box ensemble, supporting internal consistency across Andalusian health districts, though this remains a within-region check rather than external validation.

Obviously, our model is not exempt from limitations. The low precision (7.0%) and correspondingly high false discovery rate (93.0%) observed at a 60%-sensitivity operating point are an inherent consequence of screening for a low-prevalence condition (~1% in the test population) and would need to be communicated explicitly in any operational deployment, alongside the NNS framing offered above. Calibration, while modestly improved by isotonic or sigmoid post-hoc adjustment, remained imperfect at higher predicted probabilities in both calibrated and uncalibrated versions, and was not adopted for the primary classification analysis. Results for the full-variable model and for prediction horizons other than 30 days are reported only in the Supplementary Table 2, limiting the extent to which this model can characterise the trade-off between the excluded variables’ predictive contribution and the fairness rationale for their removal. Finally, despite this study was conducted entirely within a single regional health system, external validity remains to be established. Transportability may be limited by differences in EHR coding conventions and laboratory panels across health systems, and by regional differences in the epidemiological profile of undiagnosed HCV (e.g., prevalence of HIV coinfection, injection drug use). The consistent performance observed across Andalusian health districts (see Results) is reassuring against overfitting to a single local population but remains a within-region check rather than external validation. Validation in an independent health system, ideally another Spanish region, and eventually beyond Spain, is an essential next step before broader deployment.

## Data Availability

The datasets analyzed during the current study are available in the Population Health Database repository (Andalusian Regional Ministry of Health, 2021), subject to controlled access according to the regulation for the use of medical data for research in the Andalusian Health System (Andalusian Regional Ministry of Health, 2021).

## Ethical considerations

This study used pseudonymized, retrospectively extracted data obtained through the regional data-governance framework established for secondary use of health information in Andalusia (Muñoyerro-Muñiz et al., 2025). The study protocol (“Pre-HepC: Predictor of hepatitis C in undiagnosed patients based on real-world evidence”; sponsor code MRG14112022, internal code 2000-N-22, protocol version 1.0 of 14 November 2022) was reviewed and approved by the Research Ethics Committee (Comité de Ética de la Investigación, CEI) of the Hospitales Universitarios Virgen Macarena y Virgen del Rocío (Seville, Spain), which issued a favourable opinion on 20 January 2023 (meeting record CEI_01/2023). Given the retrospective, fully anonymized nature of the data extraction, the Committee granted a waiver of informed consent for the Patient Information Sheet/Informed Consent. The study was conducted in accordance with the requirements of Spanish Law 14/2007 of 3 July on Biomedical Research.

## Consent for publication

Not applicable.

## Availability of data and materials

The datasets analyzed during the current study are available in the Population Health Database repository (Andalusian Regional Ministry of Health, 2021; Muñoyerro-Muñiz et al., 2020), subject to controlled access according to the regulation for the use of medical data for research in the Andalusian Health System (Andalusian Regional Ministry of Health, 2021).

