## Supplementary for "An explainable, bias-audited machine learning model to detect hidden hepatitis C from region-wide real-world health records"

**Supplementary Table 1.** Baseline characteristics of all hepatitis C virus (HCV) cases (2017–2024), matched controls (used for training and validation) and independent random controls (used for the test set). Values are n (%) unless otherwise stated; age is expressed as mean (SD). Sex and age were matching variables for the matched controls and were not used as predictors; random controls were not matched to cases.

| Characteristic | Cases | Matched controls | Random controls |
|---|---|---|---|
| **N** | **25028** | **213404** | **193050** |
| **Sex, n (%)** | | | |
| Male | 17006 (67.9) | 145889 (68.4) | 93767 (48.6) |
| Female | 8022 (32.1) | 67515 (31.6) | 99283 (51.4) |
| Age, years, mean (SD) | 59.5 (13.3) | 59.7 (13.0) | 49.9 (17.9) |
| **Comorbidities, n (%)** | | | |
| Acquired valvular disease | 1147 (4.6) | 6875 (3.2) | 4519 (2.3) |
| Adult personality and behavior disorder | 1356 (5.4) | 2638 (1.2) | 2742 (1.4) |
| Age-related macular degeneration | 414 (1.7) | 2595 (1.2) | 2060 (1.1) |
| Alcohol dependence | 4188 (16.7) | 9019 (4.2) | 6225 (3.2) |
| Alcoholic cirrhosis of liver with ascites | 113 (0.5) | 73 (0.0) | 43 (0.0) |
| Alcoholic cirrhosis of liver without ascites | 219 (0.9) | 218 (0.1) | 102 (0.1) |
| Alcoholic fatty liver | 29 (0.1) | 82 (0.0) | 51 (0.0) |
| Alcoholic fibrosis and sclerosis of liver | 2 (0.0) | 3 (0.0) | 1 (0.0) |
| Alcoholic hepatic failure with coma | 1 (0.0) | 2 (0.0) | 1 (0.0) |
| Alcoholic hepatic failure without coma | 3 (0.0) | 9 (0.0) | 6 (0.0) |
| Alcoholic hepatitis with ascites | 7 (0.0) | 4 (0.0) | 5 (0.0) |
| Alcoholic hepatitis without ascites | 100 (0.4) | 96 (0.0) | 59 (0.0) |
| Alcoholic liver disease, unspecified | 452 (1.8) | 1001 (0.5) | 363 (0.2) |
| Aneurysm of aorta, peripheral and visceral arteries | 197 (0.8) | 1119 (0.5) | 881 (0.5) |
| Anxiety disorder | 7610 (30.4) | 45188 (21.2) | 53227 (27.6) |
| Asthma | 1854 (7.4) | 14947 (7.0) | 21250 (11.0) |
| Atopic dermatitis | 110 (0.4) | 1095 (0.5) | 4022 (2.1) |
| Atrial fibrillation | 1443 (5.8) | 9476 (4.4) | 5397 (2.8) |
| Autism spectrum disorder | 4 (0.0) | 63 (0.0) | 251 (0.1) |
| Autoimmune hepatitis | 15 (0.1) | 44 (0.0) | 54 (0.0) |
| Biliary cirrhosis, unspecified | 5 (0.0) | 13 (0.0) | 8 (0.0) |
| Bladder cancer | 280 (1.1) | 1725 (0.8) | 919 (0.5) |
| Bone and soft tissue cancer | 52 (0.2) | 251 (0.1) | 200 (0.1) |
| Breast cancer | 319 (1.3) | 2636 (1.2) | 2816 (1.5) |
| Bronchus and lung cancer | 268 (1.1) | 1077 (0.5) | 364 (0.2) |
| COPD | 3953 (15.8) | 16754 (7.9) | 9947 (5.2) |
| Cervical cancer | 74 (0.3) | 277 (0.1) | 400 (0.2) |
| Childhood and adolescence onset disorder | 385 (1.5) | 1156 (0.5) | 2561 (1.3) |

| Characteristic | Cases | Matched controls | Random controls |
|---|---|---|---|
| Chronic kidney failure | 1367 (5.5) | 5561 (2.6) | 3729 (1.9) |
| Chronic liver disease except cirrhosis | 1402 (5.6) | 984 (0.5) | 332 (0.2) |
| Chronic passive congestion of liver | 6 (0.0) | 45 (0.0) | 26 (0.0) |
| Chronic viral hepatitis B with delta-agent | 7 (0.0) | 1 (0.0) | 1 (0.0) |
| Chronic viral hepatitis B without delta-agent | 361 (1.4) | 456 (0.2) | 362 (0.2) |
| Collagen disease and vasculitis | 421 (1.7) | 2715 (1.3) | 2776 (1.4) |
| Colorectal cancer | 418 (1.7) | 2543 (1.2) | 1612 (0.8) |
| Congenital heart and circulatory anomaly | 88 (0.4) | 527 (0.2) | 669 (0.3) |
| Dementia | 693 (2.8) | 5139 (2.4) | 2419 (1.3) |
| Dependence on other substances | 3938 (15.7) | 1960 (0.9) | 2283 (1.2) |
| Diabetes | 5258 (21.0) | 37285 (17.5) | 21877 (11.3) |
| Eating disorder | 39 (0.2) | 253 (0.1) | 717 (0.4) |
| Epilepsy | 976 (3.9) | 3194 (1.5) | 2714 (1.4) |
| Extrapyramidal syndrome | 388 (1.6) | 2649 (1.2) | 2051 (1.1) |
| Fatty (change of) liver, not elsewhere classified | 1112 (4.4) | 7816 (3.7) | 6532 (3.4) |
| Fibromyalgia | 431 (1.7) | 3873 (1.8) | 4810 (2.5) |
| Gastroesophageal reflux disease | 1039 (4.2) | 9919 (4.6) | 10740 (5.6) |
| Glaucoma | 821 (3.3) | 8521 (4.0) | 5548 (2.9) |
| Gout and other crystal arthropathies | 788 (3.1) | 10048 (4.7) | 5303 (2.7) |
| HIV | 3500 (14.0) | 913 (0.4) | 566 (0.3) |
| Head and neck cancer | 262 (1.0) | 1154 (0.5) | 532 (0.3) |
| Heart failure | 2205 (8.8) | 10823 (5.1) | 5746 (3.0) |
| Hepatic fibrosis, advanced fibrosis | 0 (0.0) | 0 (0.0) | 2 (0.0) |
| Hepatic fibrosis, early fibrosis | 0 (0.0) | 0 (0.0) | 1 (0.0) |
| Hepatic fibrosis, unspecified | 10 (0.0) | 1 (0.0) | 9 (0.0) |
| Hepatic steatosis | 161 (0.6) | 449 (0.2) | 166 (0.1) |
| Hepatic veno-occlusive disease | 0 (0.0) | 0 (0.0) | 2 (0.0) |
| Hepatopulmonary syndrome | 1 (0.0) | 1 (0.0) | 0 (0.0) |
| Hepatorenal syndrome | 3 (0.0) | 9 (0.0) | 2 (0.0) |
| Hodgkin's disease | 59 (0.2) | 166 (0.1) | 133 (0.1) |
| Hypertension | 9815 (39.2) | 86785 (40.7) | 52779 (27.3) |
| Hypothyroidism | 1994 (8.0) | 16691 (7.8) | 20043 (10.4) |
| Ill-defined and other cerebrovascular disease | 233 (0.9) | 1428 (0.7) | 853 (0.4) |
| Immunoproliferative cancer | 58 (0.2) | 230 (0.1) | 141 (0.1) |
| Intellectual disability | 194 (0.8) | 1006 (0.5) | 897 (0.5) |
| Intra-abdominal arterial disease | 91 (0.4) | 378 (0.2) | 207 (0.1) |
| Ischemic heart disease | 1290 (5.2) | 11082 (5.2) | 5242 (2.7) |
| Kaposi sarcoma | 31 (0.1) | 33 (0.0) | 11 (0.0) |
| Kidney and renal pelvis cancer | 88 (0.4) | 509 (0.2) | 306 (0.2) |
| Leukemia | 97 (0.4) | 453 (0.2) | 298 (0.2) |
| Liver and biliary tract cancer | 754 (3.0) | 281 (0.1) | 101 (0.1) |
| Liver cirrhosis | 1299 (5.2) | 424 (0.2) | 89 (0.0) |
| Liver disease, unspecified | 1854 (7.4) | 1846 (0.9) | 1189 (0.6) |
| Malabsorption syndrome and food intolerance | 324 (1.3) | 2613 (1.2) | 5246 (2.7) |
| Mood disorder | 2470 (9.9) | 12658 (5.9) | 13119 (6.8) |
| Neurological disease with motor deficit, non-stroke | 1417 (5.7) | 7160 (3.4) | 5308 (2.7) |
| Non-Hodgkin lymphoma | 205 (0.8) | 838 (0.4) | 536 (0.3) |
| Nonalcoholic steatohepatitis (NASH) | 28 (0.1) | 107 (0.1) | 82 (0.0) |
| Obesity | 1709 (6.8) | 19585 (9.2) | 17541 (9.1) |
| Occlusion or stenosis of precerebral arteries | 196 (0.8) | 1435 (0.7) | 983 (0.5) |
| Osteoarthritis, spondylosis | 6794 (27.1) | 60465 (28.3) | 42175 (21.8) |
| Osteoporosis | 1479 (5.9) | 9647 (4.5) | 6625 (3.4) |

| Characteristic | Cases | Matched controls | Random controls |
|---|---|---|---|
| Other arthropathy | 1482 (5.9) | 11676 (5.5) | 9613 (5.0) |
| Other cirrhosis of liver | 87 (0.3) | 29 (0.0) | 16 (0.0) |
| Other developmental disorder | 39 (0.2) | 219 (0.1) | 669 (0.3) |
| Other functional disorder | 264 (1.1) | 948 (0.4) | 1185 (0.6) |
| Other organic mental disorder | 1769 (7.1) | 6051 (2.8) | 3961 (2.1) |
| Other specified diseases of liver | 180 (0.7) | 1172 (0.5) | 1136 (0.6) |
| Ovarian cancer | 21 (0.1) | 177 (0.1) | 212 (0.1) |
| Pancreatic cancer | 41 (0.2) | 244 (0.1) | 86 (0.0) |
| Parkinson's disease | 237 (0.9) | 1704 (0.8) | 879 (0.5) |
| Peripheral arterial disease of the limbs | 1429 (5.7) | 8615 (4.0) | 5089 (2.6) |
| Portal hypertension | 399 (1.6) | 207 (0.1) | 144 (0.1) |
| Primary biliary cirrhosis | 9 (0.0) | 55 (0.0) | 64 (0.0) |
| Prostate cancer | 231 (0.9) | 2806 (1.3) | 1642 (0.9) |
| Psoriasis | 936 (3.7) | 6791 (3.2) | 5848 (3.0) |
| Regional enteritis and ulcerative colitis | 238 (1.0) | 1750 (0.8) | 1753 (0.9) |
| Retinopathy | 1145 (4.6) | 7965 (3.7) | 5146 (2.7) |
| Rheumatoid arthritis and related diseases | 326 (1.3) | 2623 (1.2) | 2069 (1.1) |
| Schizophrenic disorder | 1445 (5.8) | 2954 (1.4) | 2048 (1.1) |
| Sequelae of cerebrovascular disease | 284 (1.1) | 1505 (0.7) | 713 (0.4) |
| Silicosis | 9 (0.0) | 44 (0.0) | 30 (0.0) |
| Skin melanoma | 63 (0.3) | 859 (0.4) | 722 (0.4) |
| Stomach cancer | 67 (0.3) | 305 (0.1) | 133 (0.1) |
| Testicular cancer | 17 (0.1) | 129 (0.1) | 111 (0.1) |
| Thyroid cancer | 38 (0.2) | 353 (0.2) | 391 (0.2) |
| Tobacco dependence | 5744 (23.0) | 24496 (11.5) | 20195 (10.5) |
| Toxic liver disease with fibrosis and cirrhosis of liver | 2 (0.0) | 0 (0.0) | 0 (0.0) |
| Unspecified cirrhosis of liver | 2073 (8.3) | 522 (0.2) | 235 (0.1) |
| Urinary lithiasis | 2427 (9.7) | 19364 (9.1) | 21464 (11.1) |
| Uterine cancer | 61 (0.2) | 490 (0.2) | 430 (0.2) |

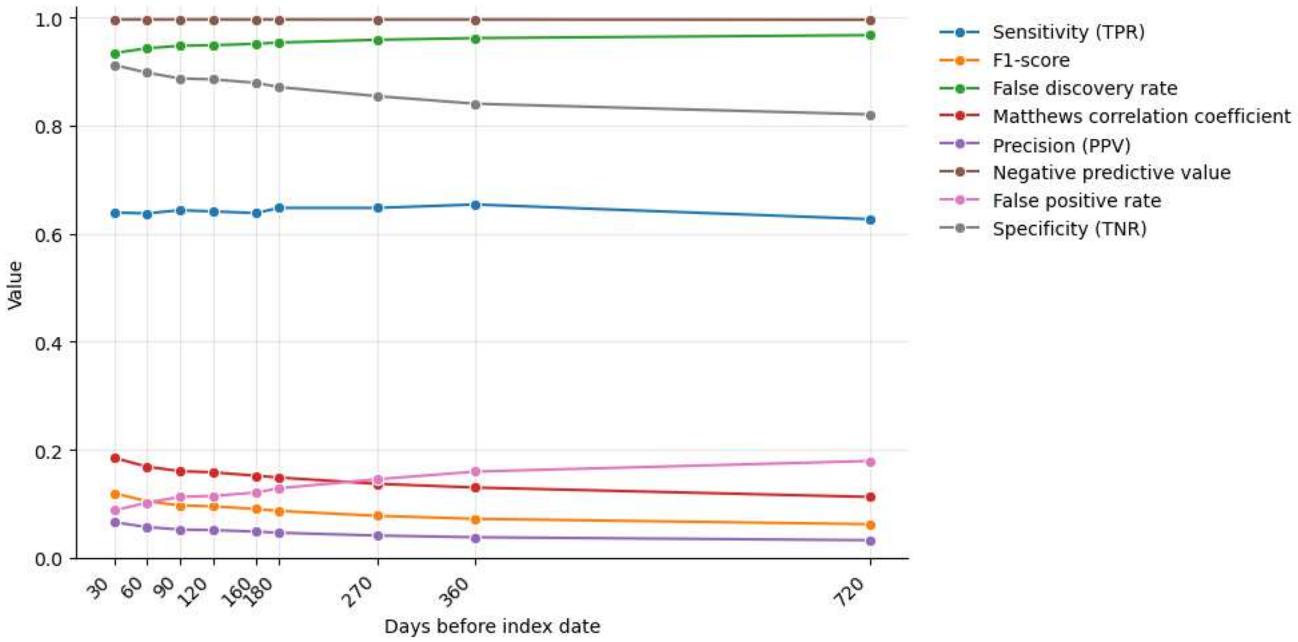


**Supplementary Figure 1.** Classification performance across prediction horizons. Sensitivity, specificity, false positive rate, positive predictive value (precision), negative predictive value, F1-score, Matthews correlation coefficient and false discovery rate in the independent test set, for models built with prediction horizons from 30 to 720 days before the index date. The classification threshold was selected in the validation set to guarantee a minimum sensitivity of 60%.

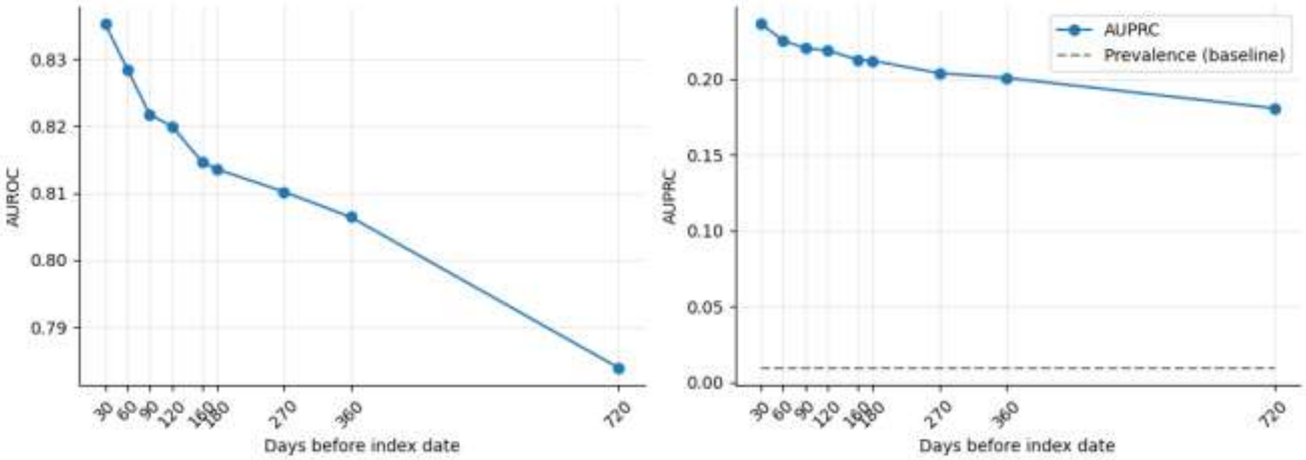


**Supplementary Figure 2.** Discrimination across prediction horizons. Area under the receiver operating characteristic curve (AUROC, left) and area under the precision-recall curve (AUPRC, right) in the independent test set for prediction horizons from 30 to 720 days before the index date. The dashed line indicates the prevalence of cases in the test set, which is the baseline AUPRC of a random classifier.

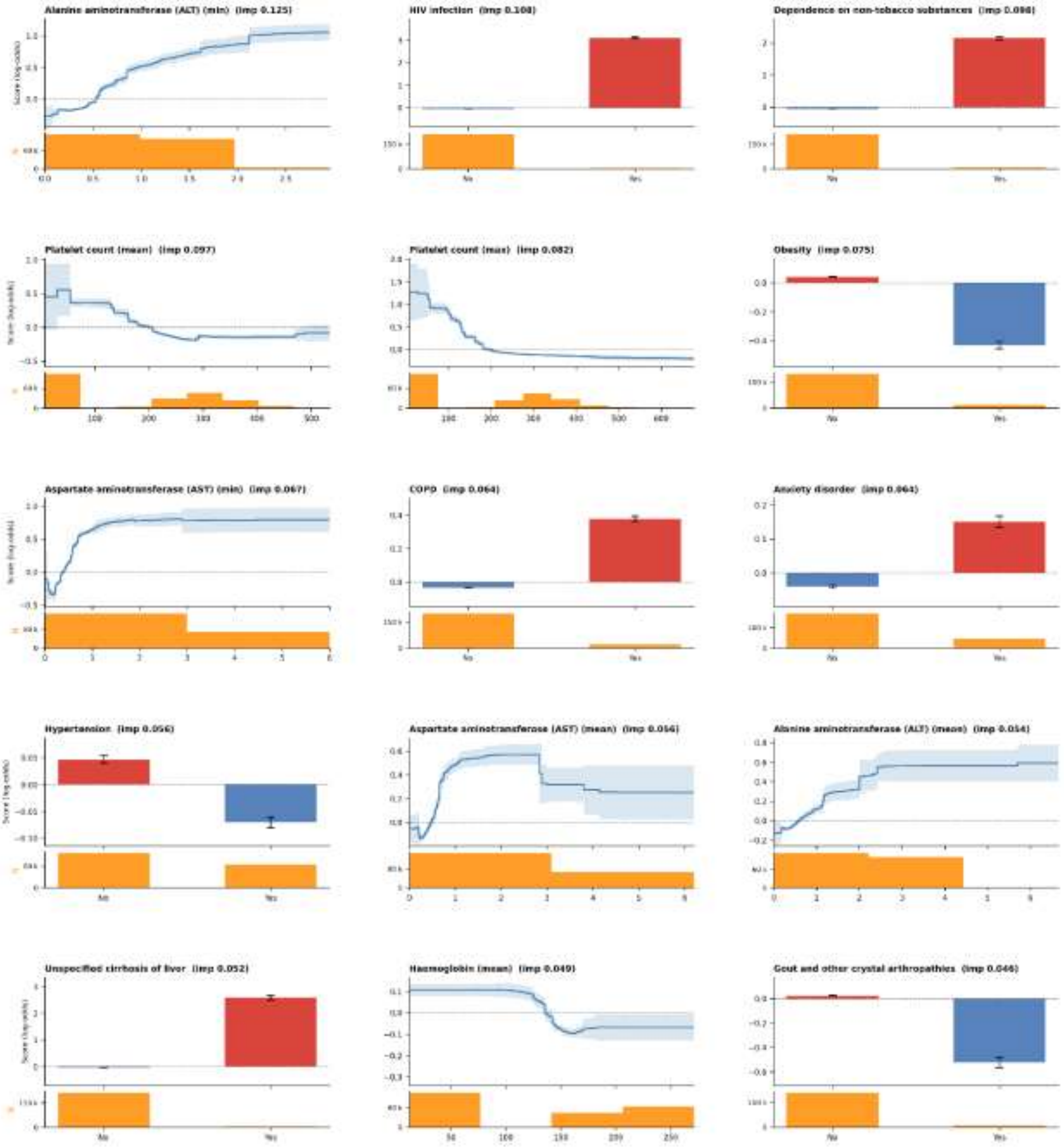


**Supplementary Figure 3.** Shape functions of the Explainable Boosting Machine for the 15 most important predictors (30-day prediction horizon). Predictors are ordered by importance (imp, mean absolute contribution to the model score). The y-axis shows the contribution to the log-odds of HCV. For continuous variables, the line is the term score and the shaded area is ±1.96 SD of the score across the bagged models of the EBM; for binary variables, bars show the score of each category with error bars of ±1.96 SD. Orange histograms show the number of individuals (N) at each value. min, mean, max and SD denote summary statistics of all laboratory determinations recorded before the prediction horizon.

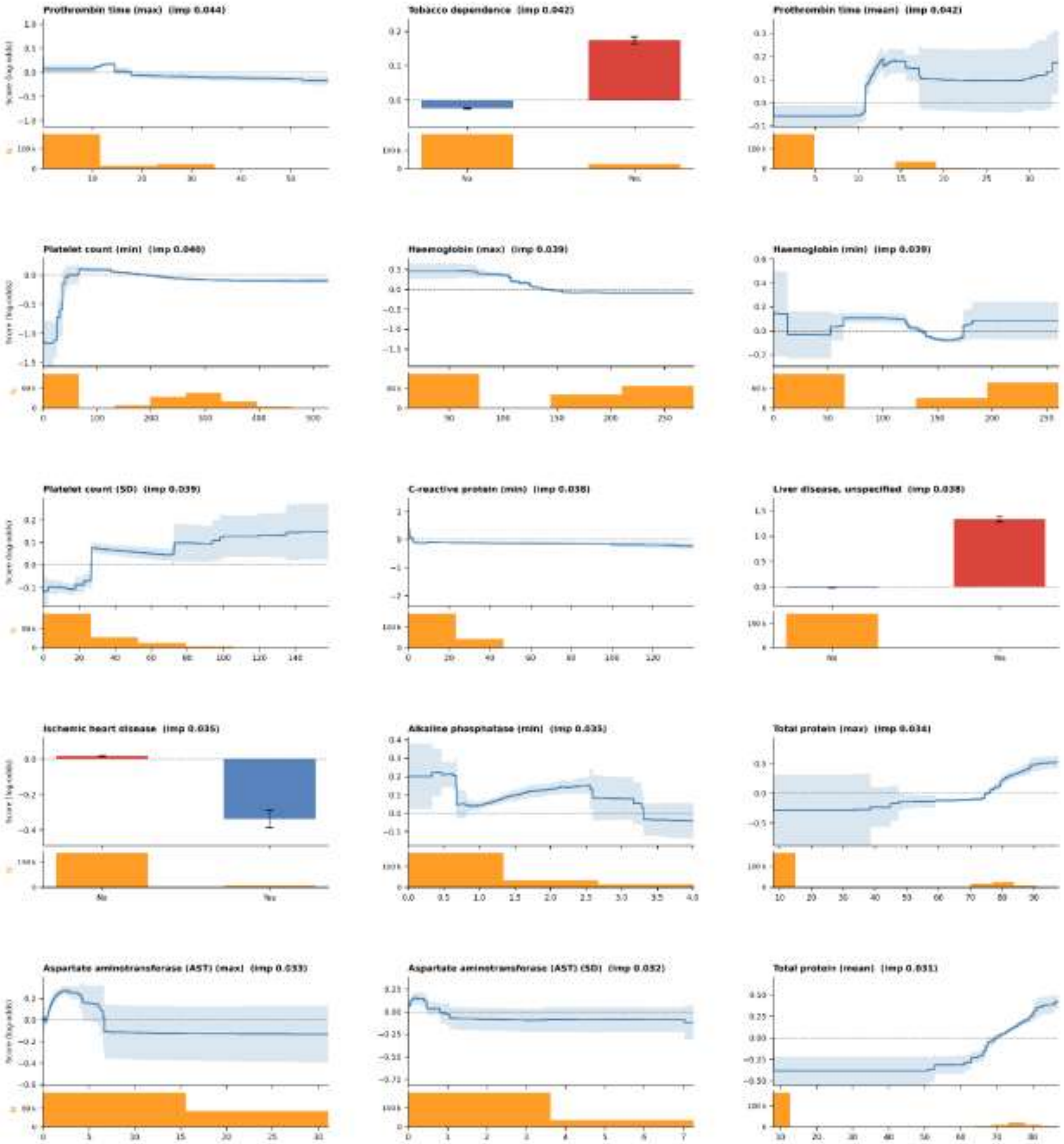


**Supplementary Figure 4.** Shape functions of the Explainable Boosting Machine for the predictors ranked 16th to 30th by importance (continuation of Supplementary Figure 3).

| Metric | Value (IC95) EBM |
|---|---|
| Sensitivity (TPR) | 0.635 (0.613–0.656) |
| Specificity (TNR) | 0.917 (0.916–0.918) |
| False positive rate | 0.083 (0.082–0.084) |
| Precision (PPV) | 0.069 (0.066–0.073) |
| Negative predictive value | 0.996 (0.996–0.996) |
| F1-score | 0.125 (0.119–0.132) |
| Matthews correlation coefficient | 0.190 (0.182–0.199) |
| False discovery rate | 0.931 (0.927–0.934) |

**Supplementary Table 2**. Classification performance of the full-variable model in the independent test set. Metrics for the Explainable Boosting Machine trained with all candidate predictors, including the income-based pharmaceutical co-payment category and the zone-level deprivation index, at the 30-day prediction horizon. The classification threshold was selected in the validation set by requiring a minimum sensitivity of 60%.
